# Understanding oral PrEP uptake and persistence among gay men and other men who have sex with men in Cape Town, South Africa: Perspectives of clients and providers

**DOI:** 10.64898/2026.11.17.26368356

**Authors:** L Long, B Brown, J Hugo, S Bokolo, C Hendrickson, C Pettinato, S Gupta, I Lakay, M Nkaphe, H Thirumurthy, MP Fox, J McIntyre

**Affiliations:** Health Economics and Epidemiology Research Office, Faculty of Health Sciences, University of the Witwatersrand, Johannesburg, South Africa; Department of Global Health, Boston University School of Public Health, Boston, MA, USA; Anova Health Institute, Johannesburg, South Africa; Department of Medical Ethics and Health Policy, University of Pennsylvania, Philadelphia, PA, USA; School of Public Health & Family Medicine, University of Cape Town, Cape Town, South Africa

**Keywords:** HIV, prevention, pre-exposure prophylaxis, men who have sex with men, South Africa

## Abstract

**Background:** Oral pre-exposure prophylaxis (PrEP) is highly effective for preventing HIV acquisition when used consistently, but uptake and persistence remain suboptimal among gay men and other men who have sex with men (MSM) in South Africa. We examined client and provider perspectives on PrEP uptake and persistence.

**Methods:** Between September and December 2022, we conducted three focus group discussions with 16 HIV-negative MSM eligible for, but not using, PrEP and six key informant interviews with PrEP providers at a specialized key population health clinic in Cape Town, South Africa. We analyzed transcripts thematically, using deductive codes informed by the PrEP cascade and the COM-B (capability, opportunity, motivation) model, plus inductive codes for emerging issues.

**Results:** Five themes emerged. Limited knowledge, misconceptions about PrEP and antiretroviral therapy, and low perceived risk reduced clients’ capability to make informed decisions about PrEP. HIV- and sexuality-related stigma, concerns about disclosure, and social surveillance were socially costly. Distance, transport costs, work commitments, clinic hours, and visit burden limited physical opportunity, whereas free care, short visits, reminders, and multi-month dispensing facilitated access. Daily pill-taking, side effects, forgetfulness, and changing sexual activity complicated persistence; routines, reminders, and acceptable PrEP modalities supported continued use. Non-judgmental, MSM-affirming services, counseling, and trusted providers fostered motivation by strengthening clients’ sense of safety, agency, and control.

**Conclusion:** PrEP delivery for MSM should extend beyond medication provision and address knowledge, stigma, service convenience, and motivation. Low-intensity adaptations, including tailored counseling, affirming care, flexible multi-month dispensing, and reminders, may improve uptake and persistence.

## Introduction

South Africa has the largest HIV epidemic in the world. In 2024, 7.8 million people were living with HIV in South Africa, of whom 6.3 million were on treatment. Life expectancy in South Africa has increased from 54.8 years (2004) to 68.3 years (2026) and the success of the antiretroviral therapy (ART) program has been largely credited for this [1, 2]. Despite these successes, South Africa still has 53,000 AIDS-related deaths each year, and HIV incidence in South Africa remains stubbornly high [3]. In 2024, there were 170,000 new HIV infections in South Africa, accounting for 13% of new infections globally [3, 4]. Highly effective biomedical HIV prevention options such as pre-exposure prophylaxis (PrEP) have the potential to significantly reduce new HIV infections [5, 6]. PrEP is one of the UNAIDS pillars of HIV prevention and is recommended for those at substantial risk of HIV infection [7]. Despite the rollout of oral PrEP in South Africa in 2016 and steadily increasing access, uptake and persistence on PrEP among the most vulnerable populations have been suboptimal [8].

Gay men and other men who have sex with men (MSM) are among the populations that are most vulnerable to HIV acquisition in South Africa [3]. The South African national strategic plan recognizes MSM as a key population for HIV prevention and supports HIV care for them through differentiated service delivery models tailored to their needs [9]. Although long-acting injectable PrEP (lenacapavir) was introduced in South Africa’s public sector in 2026 [10], oral PrEP remains the most widely available and will be a critical modality for the foreseeable future [11]. When taken consistently oral PrEP can reduce HIV acquisition by close to 90%, yet uptake and medium- to long-term persistence on PrEP among MSM and other vulnerable populations remain low globally, including in South Africa [12]. Although some discontinuation reflects genuine changes in an individual’s vulnerability, program data suggest that many clients discontinue PrEP because of challenges to continuation rather than a change in risk [8, 13]. Maximizing the impact of MSM-focused PrEP programs thus requires a detailed understanding of the specific, modifiable barriers and facilitators that shape PrEP-related behaviors, as this can allow delivery models to be adjusted to strengthen capability, opportunity, and motivation to start and stay on PrEP.

The evidence base on how best to improve PrEP care in marginalized populations remains limited, particularly in sub-Saharan Africa [14]. Early work (2012) suggested that most (75%) South African MSM were willing to use PrEP, yet this stated willingness has contrasted with low persistence in real-world settings [15, 16]. Studies from South Africa and the region have shown the roles of stigma, lack of confidentiality, perception of efficacy, pill burden, and ease of use in shaping clients’ decisions about starting and continuing PrEP [17–19]. The studies that have focused on MSM in Africa highlight this population’s own unique challenges accessing HIV and other health care services [20]. Despite this growing literature, the barriers identified to date do not fully explain why eligible individuals decline or discontinue PrEP, given its substantial health benefits. As a result, implementers have limited actionable, behaviorally informed evidence on how to adjust PrEP delivery models to encourage more optimal usage amongst those who stand to benefit the most, including MSM in South Africa.

This study was conducted to gain a better understanding of how clients and providers make decisions about using PrEP during periods of heightened vulnerability to HIV acquisition.

Human decision-making is shaped not only by structural and social constraints but also by cognitive biases and heuristics, and emerging work suggests this may also shape PrEP decisions among those most vulnerable [21, 22]. Behavioral science frameworks such as COM-B, which conceptualizes behavior as a product of capability, opportunity, and motivation, provide a structured approach to identifying aspects of PrEP service delivery that might be modifiable to address some of these behavioral barriers [23]. In this study, we used a qualitative approach (focus group discussions with MSM clients and key informant interviews with health care providers) to describe the PrEP decision-making process along the HIV prevention cascade and to classify the barriers and facilitators we identified into COM-B domains [24]. This behavioral mapping aims to generate practical, theory-driven insights into how PrEP service delivery could be adjusted to improve PrEP uptake and persistence among MSM in South Africa.

## Methods

### Overview

At a specialized key population health clinic in Cape Town (South Africa), we conducted focus group discussions (FGDs) with MSM who tested HIV negative at the clinic and key informant interviews (KIIs) with health care providers involved in the education, offering, and initiation of PrEP to clients. These data describe the PrEP decision-making process and identify how factors mapped to the COM-B domains of capability, opportunity, and motivation may contribute to poor uptake and/or persistence. We followed the COREQ (Consolidated Criteria for Reporting Qualitative Research) guidelines in reporting our methods and results [25].

### Setting

The study site is a key population clinic based in Cape Town that specializes in services for MSM and offers free services to the public. It is run by a local implementing partner that is a leader in providing specialized HIV prevention and treatment services to key populations, including MSM and transgender people. The clinic works closely with the government primary health system from which it obtains drugs, but facilities and staff are donor-funded. The clinic is situated in an urban area, is the only public clinic serving this population in the city, and so draws clients from across the city, largely individuals who cannot afford private medical insurance.

### Study design

We used a constructivist thematic analysis approach to qualitatively examine the barriers and facilitators influencing PrEP decision-making from the perspectives of clients and providers. Two frameworks guided the study. First, we organized the data collection around the HIV prevention cascade (hereafter the PrEP cascade) treating each stage, from eligibility and offer through initiation and continuation, as a distinct decision point in the client’s journey [26]. The FGD and KII guides were structured around these stages, drawing on recent work emphasizing the importance of the decision-making cascade in identifying opportunities to address behavioral barriers [27]. Second, we used the COM-B model to frame the analysis. COM-B conceptualizes behavior as the product of three interacting components: capability, opportunity and motivation [23]. Capability may be psychological (knowledge and understanding) or physical (skills); opportunity may be social (interpersonal influences and norms) or physical (environmental resources and access); and motivation may be reflective (conscious beliefs and intentions) or automatic (emotions and habits) [28]. Table 1 summarizes these domains and their potential application to PrEP use. Combining the PrEP cascade and COM-B allowed us to identify both where in the decision process barriers arose and what type of barrier they were. Including both client and provider perspectives allowed us to assess concordance and divergence in how PrEP decision-making is understood and experienced.

**Table 1.** COM-B components and their application to PrEP uptake and persistence (adapted from Michie et al. [23, 28])

| **COM-B component** | **Sub-domain** | **Definition** | **Illustrative PrEP example** |
|---|---|---|---|
| Capability | Psychological | Knowledge, understanding, and the mental skills needed to engage in the behavior | Knowing what PrEP is and how it differs from HIV treatment; understanding one’s HIV risk; remembering daily doses |
|  | Physical | Physical skills, strength, or stamina needed to perform the behavior | Physical ability to swallow pills and tolerate side effects |
| Opportunity | Social | Interpersonal influences, social cues, and cultural norms that enable or constrain the behavior | Stigma and community norms about HIV and sexuality; reactions of partners and family to PrEP use |
|  | Physical | Opportunity afforded by the environment, including time, resources, locations, and cues | Clinic location and hours, transport costs, waiting times, drug supply, dispensing intervals, and reminder cues |
| Motivation | Reflective | Conscious processes such as plans, beliefs, intentions, and evaluations | Beliefs about PrEP’s benefits and safety; seeing PrEP as protection; intentions to start or continue |
|  | Automatic | Emotions, impulses, and habits arising from associative learning | Emotional reactions such as fear or reassurance about PrEP; habitual pill-taking routines |

### Participant selection

We recruited two groups of participants: health care providers (HCPs) providing PrEP at the study site and clients eligible for PrEP who presented at the study site. KIIs and FGDs were conducted between 7 September 2022 and 8 December 2022. Hereafter, HCPs who participated in KIIs are referred to as providers, PrEP-eligible clients who participated in FGDs as clients, and both groups together as participants. Health care providers were eligible if they were actively involved in providing PrEP services at the site, had been employed for ≥6 months at the study site, and had experience in PrEP service provision for ≥6 months. They were purposively sampled to provide diversity across cadres. We approached HCPs individually, provided a brief study description, and asked whether they were interested in participating. If they were interested, we scheduled a suitable time to meet. A sample size of six for the HCPs was set to cover the majority (80%) of all PrEP-focused HCPs at the site. *PrEP-eligible clients* included all clients that reported identifying as an adult (≥18 years old) male, being HIV negative, not currently being on PrEP (at the enrollment visit), and having sex with a male within the last six months. Clients were purposively sampled to provide age diversity within and across focus group discussions. They were recruited on a day that they presented at the site for services, including an HIV test. If they tested negative, they were asked whether they were interested in participating in research and, if so, were referred to the study team. If they were eligible and still interested in participating, they were asked to return to participate in the FGD at a later date. A maximum of four focus group discussions with up to 10 participants per group was allowed for. The FGDs continued until saturation was reached. This was informed by guidance that, when focus groups and individual interviews are combined, approximately 3–4 FGDs together with 4–6 individual interviews are generally sufficient to reach saturation, and by empirical evidence that 3–6 focus groups identify around 90% of themes [29, 30].

### Data collection

For the convenience of the study participants, all study activities (KIIs and FGDs) were held in private rooms at the study site. The study team developed data collection tools based on current literature and then reviewed and refined them based on their experience. All study activities required only a single interaction with each participant. All study participants provided written informed consent, which included specific permission for audio recording. *Health care providers*: Key informant interviews with HCPs followed a semi-structured interview guide that outlined key areas and standardized prompts. The KII aimed to identify the HCPs’ role in each step of the PrEP cascade, as well as the barriers and facilitators they face in identifying suitable clients, offering PrEP, and initiating them onto PrEP. The interviewer made written notes during the KII, and the audio recording was transcribed to support the notes. Each KII was approximately 1 hour, and HCPs were offered reimbursement for their time (ZAR150 ∼ USD10). *PrEP-eligible clients:* Participants received name cards and chose a pseudonym to use during the FGDs. The discussion followed a focus group guide aimed at identifying behavioral challenges or facilitators at each stage along the PrEP cascade. The facilitator took written notes during the FGD, and the audio recording was transcribed to allow direct quotations to support the notes. Each FGD lasted approximately 1 hour, and clients received reimbursement for travel and time (ZAR200 ∼ USD13), as well as refreshments.

### Research team and reflexivity

Consistent with COREQ, we describe the research team and how our backgrounds and assumptions may have shaped data collection and interpretation [25]. LL, a researcher with training in qualitative and quantitative methods and experience conducting HIV research in South Africa, led the KIIs and FGDs. Other team members (BB, JH, JM with experience providing health care to sexual minority populations, in particular MSM) supported the development of study tools. LL had no prior relationship with participants. The team approached this work with the view that PrEP is an important public health tool and that behavioral science can improve its implementation; this framing may have led us to emphasize modifiable behavioral and service delivery factors over broader structural determinants. Throughout the study, we aimed to foreground the participants’ own experiences and situate the recommendations within the South African health system and social context.

### Data analysis

All FGDs and KIIs were transcribed from audio recordings and subsequently checked by the primary facilitator. The initial analysis was deductive, based on pre-assigned thematic codes from the study’s conceptual framework on behavioral facilitators and barriers to PrEP uptake and persistence, drawing on the COM-B model and the PrEP cascade. Two experienced reviewers (SB, CH) reviewed the transcripts using this preliminary codebook, which was then augmented using inductive code development as new themes emerged.

Next, two reviewers (CP, SG) independently coded an FGD and a KII using the evolving codebook, met to compare their coding and resolve discrepancies through discussion, and LL reviewed and refined the codebook accordingly. CP and SG then each independently coded all remaining transcripts using the final codebook in NVivo™, meeting regularly to reconcile differences by consensus. We then consolidated the final coded datasets, summarized them across reviewers, and developed themes. In the final stage of analysis, we interpreted these themes in relation to the PrEP cascade and classified them into COM-B domains to address the study aims and generate behaviorally informed implications for PrEP service delivery. To limit the influence of our own assumptions, codebooks were co-developed, coding combined deductive and inductive approaches, and local co-authors were involved in interpreting the findings.

### Ethics statement

The study was approved by the Human Research Ethics Committee (Medical) of the University of the Witwatersrand, Johannesburg, South Africa (clearance certificate M211135; approved 23 February 2022) and the Boston University Medical Campus and Boston Medical Center Institutional Review Board, Boston, MA, USA (H-40669; approved 27 July 2020). All participants provided written informed consent before taking part.

## Results

Between September 2022 and December 2022, we conducted six KIIs with providers and three FGDs with 16 clients. The providers consisted of three clinician nurses and three counselors (one working as a client liaison) with PrEP experience at the site ranging from over six months to four years. The clients all identified as male, as having had sex with another man within the last six months, ranged in age from 19 to 47 years old (mean 27), had recently tested HIV negative and indicated that they were not currently on PrEP, although they might have taken PrEP previously. We identified five overarching themes describing barriers and facilitators to PrEP uptake and persistence, presented below in relation to the COM-B model. Each participant excerpt cited below, with the complete transcript sentence(s) from which it was drawn, is provided in S1 Table.

## Theme 1. Knowledge, awareness, and misconceptions about PrEP (COM-B: Psychological Capability)

Both providers and clients indicated that limited knowledge and pervasive misconceptions about PrEP constrained uptake and persistence, as clients did not always fully understand what PrEP is, how it was meant to work, or how it differs from HIV treatment (antiretrovirals; ARVs). Confusion between PrEP and HIV treatment, questions about side effects, and often a perception of low personal HIV vulnerability meant that PrEP was viewed as unnecessary or dangerous, or as something intended only for people already living with HIV.

Across providers, there was a strong belief that clients were not fully informed about PrEP, and that this gap was amplified by community and cultural beliefs that did not align with the benefits of PrEP. One provider observed that, “*So there are people who still need education”* and that in some communities *“there are areas that they don’t even believe [in] tablets,” (KII #1)* describing a general distrust of pill-based medicine. Others supported this observation, stating, *“Yeah, because most of the people, I don’t know whether they lack education or knowledge with regards to PrEP”* (KII #2), *“I think it’s a lack of knowledge and lack of education around it”* (KII #3), and *“I think it’s lack of education.”* (KII #6)

Providers also perceived that clients misunderstood PrEP’s purpose and often conflated it with the use of ARVs for the treatment of HIV. Given the history of HIV treatment with ARVs, some clients feared that taking ARVs, even for prevention, would make them sick or cause long-term harm, and some believed that stopping PrEP in the future would result in an HIV-positive test result. As one provider noted, *“people think because of the part of ARVs they will say no they’re gonna get sick”* (KII #1), while another noted that *“there [is this] myth … if I take PrEP the next time when I stop and then I get tested I’ll be HIV positive.”* (KII #5)

Daily pill taking and the use of ARV formulations for prevention meant that for many, PrEP was equated with HIV infection rather than prevention, particularly for clients whose primary partners or family recognized pill bottles. One provider explained that *“So as long as you’re taking pills every day that’s ARVs, you’re HIV positive”* and that *“the wife will see a bottle and assume it’s ARVs”* (KII #2). Another summarized the problem succinctly: *“Bottle of tablets that means you’re HIV positive” (*KII #3).

Clients also highlighted the importance of education for their own understanding and willingness to start PrEP. Some suggested that additional explanation was needed to feel comfortable both initiating and remaining on PrEP. One client noted that *“they gave me the bottle and didn’t even explain anything about the bottle just told me [to] take one a day. So, it was so easy. Almost a bit too easy, actually”* (FGD #2). Others wanted more detailed information: *“If I didn’t know about it… explain it to me a little first before I would go on the treatment for anything”* and *“I think I would love to have like insights on why I should be on PrEP.”* (FGD #3)

Collectively, this shows how limited, and sometimes inaccurate, information about PrEP reduces psychological capability within the COM-B framework, leaving many potential clients without the basic understanding needed to appropriately consider, initiate, or sustain PrEP for HIV prevention.

## Theme 2. Stigma, identity, and social relationships (COM-B: Social Opportunity)

The social context in which PrEP is offered means there is substantial actual and perceived stigma linked to its relationship with HIV and sexuality, which in turn restricts clients’ ability to fully engage with PrEP services. Both clients and providers described how members of clients’ social networks, such as families, partners, or the wider community, often viewed clinic attendance and pill-taking as markers of HIV infection, of “*misbehaving*” (KII #4), or of sexuality. This judgment put clients’ relationships, reputations, and safety at risk, limiting social opportunities and willingness to engage with PrEP even in environments where access to services is good.

Providers commented on how key populations such as gay and bisexual men anticipated additional rejection if they were seen accessing PrEP or disclosed their sexual behavior. One provider noted that those who disclosed their sexuality were, “*neglected by families because they are gays”* and that because of *“their religion, their culture, their status… it’s very difficult for them to expose themselves in the community”* (KII #1). Another described that *“the black community they don’t like, doing this … especially with also the sexual orientation, you find that they don’t want to disclose”* (KII #6). Clients confirmed these concerns, indicating that even being *“associated with … [a] gay clinic”* and *“having other people see”* (FGD #2) was an issue.

PrEP use was also associated with infidelity or promiscuity, and some clients worried that simply possessing tablets would provoke accusations. This was particularly salient within formal relationships where clients worried “*how to hide the tablets”* (KII #1) and often felt like because they were *“married … they cannot take PrEP”* (KII #4). Even outside of marriage, PrEP use was framed negatively; providers reported that partners might “*think they’ve got multiple partners”* (KII #1) and view PrEP as evidence of “*intentions of you misbehaving”* (KII #4). Clients described similar experiences, stating that *“people are very judgmental”* (FGD #2) and that *“once you’re [on PrEP] they will think that maybe you will sleep around” and “they will think that maybe you are cheating”* (FGD #3).

Stigma extended into multigenerational households where family members monitored clients’ medications. One client shared that their “*mother… checks my medication”* (FGD #1), which contributed to a sense that they “*don’t want to leave it on the counter for everyone to see it and question it”* (FGD #2). Others worried that *“when they see it in your place… they will mistake it for maybe antiretrovirals”* (FGD #3) and therefore assume HIV infection.

These accounts show how actual and perceived stigma, moral judgment, and social surveillance at home, in relationships, and in the wider community increase the potential social costs associated with PrEP use and, as a result, reduce the social opportunity for its use.

## Theme 3. Access, convenience, and service delivery (COM-B: Physical Opportunity)

Access to PrEP was shaped by how individuals reached the site including transport time and costs as well as work schedules, waiting times, and arrangement of services at the clinic.

Clients also identified key program features, such as no cost at the point of accessing care, reminders, and multi-month dispensing, that reduced the burden of accessing PrEP, illustrating that the delivery model can either restrict or facilitate physical opportunity.

Providers were aware of the numerous structural barriers related to geography, employment, and the health system that clients faced when trying to access care, including distance to the clinic, transport expenses, and travel for work. Clients echoed these barriers. One provider observed that “*in the communities, most of them [clients] they don’t have [access] of coming this side”* (KII #4), while a client confirmed that they *“live far” and “need to take two taxis to travel”* (FGD #1) to the clinic. As a result, a major barrier is **“*…*** *financial— is transport”* (KII #1), and because clients *“are coming here by public transport*” they *“have to spend money*” to get their PrEP (FGD #3). Extended travel made consistent, timely clinic visits difficult; when providers reached out to clients, they often found people were *“out of the country, … at work”* (KII #2). For those employed, work added further logistical barriers as they *“need to be at work” and “can’t come a month later”* (KII #2), exacerbated by the fact that **“***most of them are working during the week”* while the clinic was *“not working on Saturday”* (KII #3).

At the same time, providers and clients described how free care, reduced waiting times, and strong refill systems facilitated PrEP access and use. Providers emphasized that all services at the site are offered for “*free*” (KII #1, KII #2, KII #4, KII #6), which some clients described as *“the most important”* (FGD #2) feature. Clients described the process of accessing care as “*so easy”* (FGD #2) reflecting the providers’ efforts to facilitate access by providing “*more medication so that they mustn’t default”* (KII #1) when clients were travelling, making sure “*it’s quicker to get your stuff done here than going to a public one [clinic]”* (KII #2), ensuring that *“we never run out of stock”* (KII #2), and organizing visits so that *“it doesn’t take that much of their time”* (KII #5). Clients corroborated these efforts, reporting multi-month dispensing if *“I won’t be back”* (FGD #1) and having clear appointment reminders, such as *“on my bottle they wrote that I need to come back on the 19 of December”* (FGD #3).

Consistently, these accounts show that structural factors such as distance, transport costs, work schedules, and health-system organization limit the physical opportunity for PrEP use, while free care, efficient service delivery, reminders, and multi-month dispensing can partially offset these barriers within the COM-B framework.

## Theme 4. Pill burden, side effects, and adherence practices (COM-B: Psychological Capability, Physical Opportunity, and Automatic Motivation)

While initiation of PrEP required a willingness and recognition of need, persistence required clients dealing with the day-to-day work of taking a pill every day, managing side effects, and adding PrEP into their current schedules. Clients shared how pill burden, adverse effects, forgetfulness, and fluctuating sexual activity resulted in unintended breaks or discontinuation, while establishing habits or using reminders helped some remain engaged. Preferences for alternative future PrEP modalities (e.g., implants, long-acting injections) reflected attempts to address these obstacles.

Providers and clients flagged that daily pill-taking, forgetting doses, experiencing side effects, and changes in sexual activity all influenced adherence and persistence. One provider shared that *“people … are tired of taking PrEP drugs, the pill, each and every day”* (KII #2) and that this was amplified by the cognitive dissonance clients experience of *“taking a pill every day yet I’m not sick”* (KII #6). For others, providers felt the barrier was less about the pill itself than about remembering it *“… because they always forget sometimes that I had to take my tablet”* (KII #1), a challenge echoed in focus groups where clients acknowledged *“forgetting to drink it every day”* (FGD #3). Although side effects were generally mild and temporary, providers reported that some clients did not feel comfortable taking the medication *“because of the side effects”* (KII #4), which included “*headaches*” or “*nausea*” and led some to conclude that they “*… don’t like these tablets, then they leave them*” (KII #1). Clients likewise reported that *“PrEP makes me feel not good”* (FGD #1) and one client described friends on PrEP who had experienced “*weight gaining*”, “*feeling tired*”, and “*feel[ing] depressed*” (FGD #1). For some, decisions to stop were framed primarily in terms of perceived risk, with providers recounting clients who stopped because they were “*not sexually active*” (KII #2, KII #5) and others who took PrEP because they were “*sexually active*” and had “*more partners*” (KII #1).

In acknowledging challenges, clients also described practical adherence strategies they had adopted. These included phone alarms (“*If you are starting … you put an alarm*”, FGD #3), visual cues tied to existing routines (“*I just put it in [a] place that I can see it frequently*”, FGD #2), and external support from partners (“*he will remind me in the morning*”, FGD #2).

Clients saw these strategies as ways of “*getting into a routine*” (FGD #3) that could support consistent adherence over time. When asked about future potential PrEP modalities, clients framed their preferences as strategies for improving adherence and persistence, though preferences were not consistent across options, illustrating that different modalities can either support or undermine adherence depending on context and individual. One client expressed a preference for “*an implant because I for one I hate pills*” (FGD #3), while another stated that they *“enjoy drinking pills in the morning”* because it gave them the feeling that they were *“in control”* (FGD #3). Similarly, one client thought a *“once-off injection per month it will be better”* (FGD #3), but others preferred the pill because they felt that with injections *“there’s so many more things that could go wrong”* (FGD #2).

These narratives highlight how pill burden, side effects, forgetfulness, and changes in sexual activity can reduce both psychological capability and physical opportunity for sustained PrEP use, while habitual routines (automatic motivation), reminder cues, and acceptable PrEP modalities (physical opportunity) helped some clients sustain PrEP use.

## Theme 5. Counseling, psychosocial comfort, trust and motivation for protection (COM-B: Reflective Motivation and Social Opportunity)

Both providers and clients reported that accessible counseling, peer-centered communication, and welcoming and affirming clinical spaces enabled clients to communicate directly about sexual health, understand HIV prevention options, and identify themselves as active participants in the protection of their health. These interpersonal and structural features improved reflective motivation (desire to “*take care of myself*” [KII #6, describing clients’ views], “*stay HIV negative” [FGD #1])* and social opportunity (non-judgmental clinical spaces).

Providers noted that clear, informative counseling that explained the benefits, risks, and guidelines helped clients to “*understand themselves*” (KII #5), recognize risk, and make appropriate decisions about PrEP. Providers noted that, “*you need to show the person or teach the person*” (KII #1); that once you have “*educated them well*” they “*understand the benefits of taking PrEP*” (KII #6), which allows them to be “*responsible*” and “*aware of your risk behavior*” (KII #5). They actively made supportive space for clients to come to their own decisions by providing those not ready with “*small pamphlets for PrEP*” and they “*write on the pamphlets our cell phone numbers… so if ever you change your mind, you can just give us a call*” (KII #3).

Providers emphasized the role that better clinical environments (friendly, men-focused, gay-friendly, and non-judgmental) can play in facilitating open conversation on sensitive topics (sexual orientation, partners, drug use), and clients described their experience of the study clinic as feeling “*relieved*” (FGD #1) and “*included*” (FGD #2), often in contrast to their home and community settings which were less welcoming. The fact that clients could “*just see it’s gay friendly*” (KII #3) and that providers were perceived as “*easily approachable*” (KII #1) resulted in providers being able to get clients to “*engage, feel comfortable, and then you’ll get all the answers you want*” (KII #2). Clients described relief at finding a place where “*they specialize in men’s health*” (FGD #2) and “*you feel like no one is judging you*” and “*have all the information that you’re looking for…*” (FGD #1). Part of the reason for feeling included and motivated was that clients described having providers who were “*experiencing the same lifestyle that I experienced…*” (FGD #2) and that looking around the clinic it was “*good just to see other men in our community taking care of themselves*” (FGD #2). The benefits of such a setting therefore need to be considered together with the knowledge gaps, stigma, and practical barriers described in the other themes.

Clients across all the FGDs framed accessing health care, including PrEP, as part of a broader prevention mindset, linking services like STI screening, counseling, and PrEP with positive mental health and feeling “*in control*” (FGD #3). Clients described PrEP as a “*safety net*” to help stay HIV negative when the unexpected happens (“*a condom breaks*”) or when their personal situation (multiple partners, unknown partner status) places them at risk, allowing them to “*live your life and enjoy*” and giving a “*peace of mind that comes with knowing that you are safe*” (FGD #1). “*HIV testing and counseling*” (FGD #3) were given as one of the main reasons for coming to the clinic, giving clients “*more control*” (FGD #2). The habit of “*booking a routine checkup every three months*” (FGD #2) encouraged not only PrEP persistence but also regular STI screening (“*kept me in line with like getting STI screen*”, FGD #1).

Together, these accounts suggest the combined role of quality counseling delivered by friendly community providers in a welcoming clinical environment in promoting reflective motivation and social opportunity for PrEP, building clients’ agency for risk identification and management, which clients linked to a broader sense of taking care of their health.

## Discussion

In interviews and discussions with providers and MSM in South Africa, we found that PrEP use was shaped by multiple decisions along the PrEP decision cascade from initiation to persistence that fall into distinct domains in the COM-B framework for studying behavior change. These include lack of knowledge and pervasive misconceptions (psychological capability), stigma and social surveillance (social opportunity), service delivery constraints (physical opportunity), pill burden and adherence challenges (psychological capability, physical opportunity, and automatic motivation), and the clinical environment and counseling (reflective motivation and social opportunity). Taken together, these findings help explain the continuing gap between high reported willingness to use PrEP and low real-world uptake and persistence amongst MSM and other vulnerable populations in South Africa [31].

Despite counseling and education efforts, there remained gaps in knowledge around PrEP use that allowed myths and misconceptions to play substantive roles in decision-making (e.g., PrEP = ARVs, which is for sick people, but I am not sick; stopping PrEP will turn tests positive). These factors reduce the psychological capability of individuals to evaluate PrEP initiation or persistence, and similar patterns have been reported in adolescent girls and young women in Malawi [32] and elsewhere in sub-Saharan Africa, as well as in other vulnerable populations in South Africa [33].

For those who do start PrEP, there remain barriers outside of physically accessing and taking the medication. PrEP can act as an undesirable social marker, interpreted as living with HIV, being promiscuous, or being gay, each of which can trigger judgment from partners, families, and communities. This dynamic constrains social opportunity, and even specialized MSM clinics can themselves be stigmatizing if attendance is visible or recognizable. These results are consistent with prior work among MSM and other key populations in South Africa, which has identified stigma, privacy concerns, and the reactions of clients’ social networks as central barriers to PrEP uptake and persistence [31, 33].

Service design (including clinic location, operating hours, waiting times, and visit frequency) also shaped the physical feasibility of accessing PrEP services; participants highlighted multi-month dispensing, appointment reminders, and free services as particularly valued features. Such service delivery factors largely determine physical opportunity and have been shown to influence PrEP persistence in many South African facilities [34]. Our findings suggest that misunderstandings about PrEP use and safety may act upstream in the decision cascade, preventing clients from self-identifying as suitable PrEP candidates or accepting an initial offer, while stigma and social surveillance are more likely to become salient downstream, when clients must decide whether to store tablets at home or return to the clinic for a refill. Lastly, service design likely acts both upstream and downstream, affecting clients’ first encounters with the health system as well as their experience of continued PrEP use.

Participants’ accounts suggest that MSM-affirming, non-judgmental clinic settings may improve motivation to access care in the first place and support persistence once PrEP was initiated by creating an environment of trust and perceived control. Similar patterns have been reported in prior work demonstrating that patient-centered, stigma-free services are associated with improved engagement in HIV prevention and care among key populations [14, 19]. Clients in our study who viewed PrEP as a way to proactively protect themselves and take control of their health described greater motivation to use PrEP. This aligns with the literature showing that self-efficacy and positive affect are key determinants of adherence to both antiretroviral and PrEP programs [35–38]. Conversely, when PrEP was framed as evidence of sickness rather than proactive protection, it undermined clients’ motivation to consider PrEP initiation, and further along the cascade, continued exposure to stigma coupled with pill fatigue eroded motivation to continue. This observation aligns with behavioral frameworks like COM-B, which highlight that reflective motivation (beliefs and intentions) and automatic motivation (emotional responses) are not static and can change over time in response to clinical and social context [39]. These findings add to South African and global MSM PrEP studies that highlight the importance of tailored counseling, staff training, and clinic design in overcoming fear, mistrust, and stigma when accessing healthcare. Our COM-B-informed analysis therefore not only helps explain why some MSM may initiate and persist on PrEP while others do not but also points to practical levers for improving PrEP services in resource-constrained settings.

Our findings point to several behaviorally grounded interventions for improving PrEP service delivery and the health of individuals at risk of HIV acquisition. First, PrEP counseling should go beyond information provision to build skills: correcting common myths and misinformation about PrEP and supporting clients to accurately and independently assess their own risk.

Counseling can also foster identity-based motivation, helping clients see themselves as people who take care of their health and communities, which participants linked to peace of mind and persistence.

Second, programs should actively address the social costs of taking PrEP by providing practical guidance on pill storage and packaging to reduce the likelihood that tablets are interpreted as HIV treatment or evidence of infidelity, and by offering optional couple or family sessions and simple communication scripts to help clients negotiate disclosure in ways that protect relationships and promote safety. Third, programs should prioritize service design features that enhance physical opportunity (multi-month dispensing, flexible clinic hours, shorter visits), which directly address the logistical barriers clients described and make it easier to build stable PrEP-taking routines (proactive SMS or app-based reminders for pill and visit adherence) around work and life demands.

Finally, clients placed a high value on MSM-affirming branding, staff from and familiar with their community, and consistently non-judgmental interactions, and these elements should be recognized as core components of effective PrEP delivery programs, not optional extras. Programs can invest in ongoing staff training, strong community linkages, and visible signals of safety (e.g., inclusive posters and messaging, “safe provider” designations) to reinforce trust and motivation across the cascade, including in general public sector facilities that will need to serve these populations. Within the COM-B framework, this evidence suggests that light-touch interventions – such as decision aids that reframe and demedicalize PrEP as protection, default policies favoring extended multi-month dispensing, and built-in reminder systems – could support both uptake and persistence among MSM. These light-touch interventions are particularly important in the current funding environment, where dedicated resources for vulnerable populations are being reduced. This has pushed these populations out of functional clinical environments into fragmented public clinical systems that are not always well equipped or trained to meet the needs of MSM and other vulnerable populations.

This study has several strengths. It integrates both provider and client perspectives from the same key population clinic, allowing us to compare how clients and providers view the same barriers and facilitators, and it applied a COM-B-informed decision cascade to PrEP uptake and persistence, providing a structured, theory-driven lens for unpacking behavioral challenges in this context. However, limitations of this study include the focus on a single, urban, donor-supported key population clinic. Our findings may thus not fully generalize to public sector or rural facilities. In addition, clients were recruited at the clinic and were therefore already engaged with health services; the perspectives of MSM who do not access care may differ. Data were also collected before recent reductions in dedicated services for vulnerable populations, including MSM, in South Africa, which may further exacerbate the barriers described here [40, 41]. Future research should assess the effectiveness of behaviorally informed modifications to PrEP delivery at scale – for example, structured counseling, service design adjustments, and reminder systems – to determine how well they improve key pathways to behavior. It will also be important to extend this foundational work to other vulnerable populations and modalities (e.g., long-acting injectable lenacapavir, now being rolled out in South Africa) to understand whether the mechanisms driving behavior are shared or population- and context-specific, particularly as PrEP delivery moves into more generalized care settings.

Using a prominent framework for studying behavior change, this study highlights that PrEP success among MSM depends not only on access to medication, but on navigating the pathways to PrEP uptake and persistence through the ways information, stigma, clinic environment and service design shape capability, opportunity and motivation across the decision cascade. Harnessing these behavioral insights through light-touch, scalable interventions will be increasingly important if PrEP programs are to remain effective amid shrinking resources and growing reliance on public-sector services.

## Author Contributions

Conceptualization – LL, BB, JH, JM Data Curation - LL Analysis – LL, CP, SG, CH, SB Funding Acquisition - LL Investigation – LL, BB, JH, IL, MN Methodology – LL, MF, HT, JM Writing – Original draft preparation - LL Writing – Review and editing - All

## Data Availability

The data underlying this study are qualitative transcripts from focus group discussions with men who have sex with men and key informant interviews with healthcare providers in South Africa. Participants were informed that transcripts and recordings would be accessible only to study staff and that results would be reported without attribution to identifiable individuals they did not consent to public sharing of full transcripts. Given the small number of participants, the sensitivity of the topics (sexual behavior, HIV prevention), and the risk of indirect identification, the full transcripts cannot be shared publicly. All de-identified excerpts supporting the findings are included in the manuscript and in S1 Table. Requests for access to additional de-identified data may be directed to the Human Research Ethics Committee (Medical), University of the Witwatersrand (Tel: +27 11 717 2700), citing protocol M211135, and will be considered subject to ethics approval and participant consent conditions.

## Acknowledgements

The authors gratefully acknowledge the clients and health care providers who shared their time, experiences, and perspectives. The authors also thank the clinic and study-site staff for supporting study implementation, participant engagement, and data collection. We are grateful to the mentors, collaborators, and advisory team supporting the NIMH K01 award for their scientific guidance, mentorship, and constructive feedback throughout the development of this work.

## Competing and Conflicting Interests

No competing or conflicting interests were disclosed.

## Data Availability Statement

The data underlying this study are qualitative transcripts from focus group discussions with men who have sex with men and key informant interviews with healthcare providers in South Africa. Participants were informed that transcripts and recordings would be accessible only to study staff and that results would be reported without attribution to identifiable individuals; they did not consent to public sharing of full transcripts. Given the small number of participants, the sensitivity of the topics (sexual behavior, HIV prevention), and the risk of indirect identification, the full transcripts cannot be shared publicly. All de-identified excerpts supporting the findings are included in the manuscript and in S1 Table. Requests for access to additional de-identified data may be directed to the Human Research Ethics Committee (Medical), University of the Witwatersrand (Tel: +27 11 717 2700; Email), citing protocol M211135, and will be considered subject to ethics approval and participant consent conditions.

## Supporting information

S1 Table. Participant excerpts cited in the manuscript, with the complete transcript sentence(s) from which each excerpt was drawn. Excerpts are numbered in order of appearance in the Results. Full sentences are reproduced verbatim from de-identified transcripts; square brackets mark transcriber notes, redactions, or words omitted to protect participant confidentiality. KII = key informant interview (provider); FGD = focus group discussion (client). (XLSX)

## Notes

### Competing Interest Statement

The authors have declared no competing interest.

### Author Declarations

The study was approved by the Human Research Ethics Committee (Medical) of the University of the Witwatersrand, Johannesburg, South Africa (clearance certificate M211135 approved 23 February 2022) and the Boston University Medical Campus and Boston Medical Center Institutional Review Board, Boston, MA, USA (H-40669 approved 27 July 2020). All participants provided written informed consent before taking part.

